# Between-networks hyperconnectivity is induced by beta-amyloid and may facilitate tau spread

**DOI:** 10.1101/2024.01.03.24300709

**Authors:** Seyed Hani Hojjati, Tracy A. Butler, Mony de Leon, Ajay Gupta, Siddharth Nayak, José A. Luchsinger, Gloria C. Chiang, Qolamreza R. Razlighi

**Affiliations:** Department of Radiology, Brain Health Imaging Institute, Weill Cornell Medicine, New York, NY, United States; Department of Radiology, Quantitative Neuroimaging Laboratory, Brain Health Imaging Institute, Weill Cornell Medicine, New York, NY, United States; Departments of Medicine and Epidemiology, Columbia University Irving Medical Center, New York, NY, United States

**Keywords:** Between-networks hyperconnectivity, tau, beta-amyloid, positron emission tomography (PET) and resting-state functional magnetic resonance imaging (rs-fMRI)

## Abstract

Alzheimer’s disease (AD) is characterized by the buildup of neurofibrillary tau tangles and beta-amyloid (Aβ) plaques. While it has been hypothesized that Aβ facilitates the spread of tau outside of the medial temporal lobe (MTL), the specific pathological processes and mechanisms by which this occurs remain poorly understood. Our study employed advanced neuroimaging techniques, integrating 18F-Florbetaben Aβ and 18F-MK6240 tau positron emission tomography (PET) with resting-state functional magnetic resonance imaging (rs-fMRI) to characterize these mechanisms in two distinct datasets, that included 481 healthy elderly subjects, 46 of whom came with longitudinal data. Our research highlighted an intricate internetwork relationship between Aβ and tau accumulation, across spatially distinct functional networks. Additionally, we observed compelling evidence supporting the existence of a compensatory mechanism triggered by Aβ accumulation, resulting in hyperconnectivity between functional networks. Finally, the longitudinal findings indicate that between-networks hyperconnectivity is associated with future tau elevation and mediates the relationship between cortical Aβ and early-stage tau. Understanding this early brain alteration in response to the accumulation of Aβ could guide treatments early in the disease course and potentially prevent future tau accumulation.

## 1. INTRODUCTION

Alzheimer’s disease (AD) is a prevalent neurodegenerative condition primarily affecting the elderly. It is a progressive disorder that leads to cognitive decline, memory loss, and changes in behavior^1–4^. The disease is characterized by two distinct types of pathological changes in the brain: neurofibrillary tau tangles inside neurons and the accumulation of beta-amyloid (Aβ) plaques outside neurons^5–7^. Previous studies have explored several mechanisms by which tau pathology develops and spreads, such as the abnormal transmission of signals between nerve cells, excitotoxicity, and the prion-like spread of pathogenic misfolded proteins^7^. More importantly, it has been hypothesized that the spread of tau out of the medial temporal lobe (MTL) is facilitated by Aβ^5,8,9^. However, the exact mechanism behind this facilitation is still a subject of debate among researchers. The main challenge in understanding this hypothesis revolves around the distinctive spatial initiation and, to some extent, progression of Aβ and tau pathologies^5,10^. Several recent imaging studies have demonstrated that the brain’s functional connectivity plays a pivotal role in the observed patterns of tau deposition^11–13^. The spread of tau in more functionally connected regions may serve as a pathway for explaining the broader associations between Aβ in remote regions, facilitated by neuronal projections. Therefore, the central premise of our study rests on how functional connectivity might mediate the remote associations between Aβ and tau in the early stages of accumulation.

Most research on Aβ toxicity has primarily focused on its deteriorative effects, leading to significant reductions in synaptic activity and neuronal metabolism^14,15^. However, the impact of Aβ on neural function, which may transiently rise early in the disease process through compensatory responses^16^, remains largely unexplored. In addition, AD research has predominantly focused on the impact of Aβ pathology on alterations in within-network functional connectivity^17–19^, which examines the connections and interactions within specific brain networks. However, the investigation of how Aβ may affect connections between different functional networks has been less explored. Recent evidence has reported an observable reduction occurs in the specificity of functional networks and integration between them during normal aging^20,21^. These alterations in brain functional connectivity may signify an adaptive compensatory response aimed at mitigating the effects of Aβ pathology, resulting in increased connectivity between certain functional networks. This can provide insights into how Aβ pathology influences the broader communication and coordination among distinct brain regions or functional networks^22–25^. Finally, this neural connection increase can lead to an increase or even spread of tau pathology^11,12,26^.

Utilizing multimodal neuroimaging, Aβ positron emission tomography (PET), and tau PET imaging with resting-state functional magnetic resonance imaging (rs-fMRI), has significantly enhanced our ability to explore the intricate relationship between brain connectivity and pathology. We hypothesize that the brain undergoes changes in neural function, specifically between-networks hyperconnectivity, through compensatory mechanisms in response to Aβ aggregation years before clinical symptoms manifest. This underlying mechanism may explain how Aβ can trigger the spread of tau even across spatially distinct brain regions and functional networks. To achieve our research objectives, we validated our hypotheses through comprehensive statistical analyses and the application of graph theory measurement. This innovative research has the potential to offer a comprehensive view of the underlying mechanisms involved in the early-stage interplay between Aβ and tau pathologies. Furthermore, this research can potentially lead to development of early biomarkers for AD.

## 2. METHODS

### 2.1. Sample characteristics

We included 361 healthy control (HC) elderly individuals from the Northern Manhattan Study of Metabolism and Mind (NOMEM) [mean age 64.96 ± 3.17 years, 230 females] as part of the ongoing study at Columbia University Irving Medical Center^27^. Forty-six subjects in this dataset have follow-up scans (mean age 64.23 ± 3.15 years, 26 females) with data acquisition intervals ranging from 2 to 3 years. We also included 120 HC elderly individuals from the Brain Health Imaging Institute (BHII) [mean age 68.71 ± 6.16 years, 54 females] studies conducted at Weill Cornell Medicine. All participants underwent MRI, 18F-Florbetaben, and 18F-MK6240 PET imaging (see the next section). We also used younger subjects (**Table S1**) who had either a tau-PET scan (47 subjects, mean age 29.36 ± 4.73 years, and 27 females) or an Aβ-PET scan (97 subjects, mean age 27.64 ± 3.23 years, and 55 females) as part of the harmonized PET scan protocols. These young, healthy subjects served as a normative reference group, allowing us to generate region-specific and global cut-points^28,29^. All individuals underwent comprehensive evaluations that encompassing both medical and neuropsychological assessments. These assessments confirmed the absence of neurological or psychiatric disorders, cognitive impairment, and significant medical conditions. Our data have undergone thorough ethical approval through the processes of two institutional review boards (IRBs), located at Columbia University Irving Medical Center and Weill Cornell Medicine. Moreover, informed consent was obtained from all individuals for all experiments and neuroimaging scans. **Table 1** provides a complete overview of each group of subjects’ demographics, pathological status, and cognitive scores.

**Table1.** Demographics and clinical characteristics of the population in this study.

|  | NOMEM<br>(n=361) | BHII<br>(n=120) | Statistics |
| --- | --- | --- | --- |
| Age | 64.96 (3.17) | 68.71 (6.16) | P<0001 |
| Sex (M / F) | 131 / 230 | 66 / 54 | P<0001 |
| Race (Hispanic / Non-Hispanic) | 256 / 105 | 0/120 | P<0001 |
| Number of subjects with follow-ups | 46 | 0 | - |
| Global Aβ SUVR | 1.21 (0.16) | 1.24 (0.33) | P>0.26 |
| BRAAK stages I-II tau SUVR | 0.96 (0.16) | 0.97 (0.15) | P>0.45 |
| BRAAK stages I-VI tau SUVR | 0.98 (0.26) | 1.00 (0.24) | P>0.46 |
| Category Fluency | 43.82 (10.85) | 39.21 (9.77) | P<0.01 |
| Selective Reminding Test – LTS score | 31.04 (13.79) | - | - |
| Selective Reminding Test – CLTR score | 21.43 (14.14) | - | - |
| Selective Reminding Test – SRT: Delayed Recognition score | 10.78 (1.94) | - | - |
| Selective Reminding Test – SRT: Delayed Recall score | 5.67 (2.45) | - | - |
| Blind MOCA score | - | 19.57 (2.11) | - |
| RAVLT Immediate score | - | 44.75 (9.52) | - |
| RAVLT Learning score | - | 6.47 (2.09) | - |
| RAVLT Forgetting score | - | 2.81 (1.79) | - |

| NOMEM dataset | nAβ/nTau<br>(n=242) | aAβ/nTau<br>(n=39) | nAβ/aTau<br>(n=54) | aAβ/aTau<br>(n=26) | P-value |
| --- | --- | --- | --- | --- | --- |
| Age | 64.73 (3.05) | 65.35 (3.05) | 65.22 (3.59) | 66.00 (3.18) | P>0.17 |
| Sex (M/F) | 90 / 152 | 15 / 24 | 19 / 35 | 7 / 19 | P>0.75 |
| Global Aβ SUVR | 1.15 (0.05) | 1.37 (0.14) | 1.16 (0.05) | 1.60 (0.31) | P<0.0001 |
| BRAAK Stage I-II tau SUVR | 0.88 (0.10) | 0.87 (0.11) | 1.62 (0.16) | 1.60 (0.50) | P<0.0001 |
| BRAAK stages I-VI tau SUVR | 0.91 (0.09) | 0.89 (0.08) | 1.09 (0.09) | 1.29 (0.25) | P<0.0001 |

| BHII dataset | nAβ/nTau<br>(n=73) | aAβ/nTau<br>(n=17) | nAβ/aTau<br>(n=19) | aAβ/aTau<br>(n=11) | P-value |
| --- | --- | --- | --- | --- | --- |
| Age | 68.38 (5.41) | 67.35 (5.59) | 66.89 (5.46) | 66.54 (4.35) | P<0.001 |
| Sex (M / F) | 37 / 36 | 11 / 6 | 11 / 8 | 7 / 4 | P>0.66 |
| Global Aβ SUVR | 1.09 (0.05) | 1.63 (0.23) | 1.067 (0.06) | 1.93 (0.42) | P<0.0001 |
| BRAAK stage I-II tau SUVR | 0.90 (0.09) | 0.92 (0.10) | 1.14 (0.1) | 1.51 (0.39) | P<0.0001 |
| BRAAK stages I-VI tau SUVR | 0.91 (0.07) | 0.92 (0.08) | 1.07 (0.05) | 1.27 (0.25) | P<0.0001 |

NOMEM: Northern Manhattan Study of Metabolism and Mind; BHII: Brain Health Imaging Institute; Aβ : Beta-amyloid; M: Male; F: Female; SUVR: Standardized uptake value ratio (SUVR); LST: Long-term storage; CLTR: Consistent long-term retrieval; SRT: Short-term recall; MOCA: Montreal cognitive assessment; RAVLT: Rey auditory verbal learning test; nAβ /nTau: Normal Aβ and normal tau; aAβ /nTau: Abnormal Aβ and normal tau; nAβ /aTau: Normal Aβ and abnormal tau; aAβ /aTau: Abnormal Aβ and abnormal tau.

### 2.2. Image acquisition

All MRI scans were conducted using 3.0 Tesla MRI scanners. Each participant’s imaging session began with a scout localizer to determine positioning and field of view, followed by a high-resolution magnetization-prepared rapid gradient-echo scan. In the NOMEM dataset, the MRI was performed with a TR/TE (time of repetition/time of echo) of 7/2.6 ms, a flip angle of 12°, a matrix size of 256×256, and 176 slices with a thickness of 1 mm. In the BHII dataset, the MRI was performed with a TR/TE of 2400/3 ms, a flip angle of 9°, a matrix size of 512×512, and 416 slices with a thickness of 0.5 mm.

In the NOMEM dataset, resting-state fMRI was performed with a TR/TE of 2000/23 ms, flip angle of 77°, matrix size of 128×128, voxel size of 1.5×1.5×3 mm, and 40 axial slices, each lasting 10 minutes. In the BHII dataset, resting-state fMRI was performed with a TR/TE of 1008/37 ms, flip angle of 52°, matrix size of 104×104, voxel size of 2×2×2 mm, and 72 axial slices, each also lasting 10 minutes.

Both of this study’s datasets followed a standardized procedure for acquiring Aβ- and tau-PET images. For tau-PET imaging, all participants received the 18F-MK6240 tracer. The tracer was introduced via an intravenous catheter in the arm, with an injection of 185 MBq (5 mCi) ± 20% (maximum volume 10 mL), administered as a single IV bolus within 60 seconds or less (equivalent to 6 secs/mL max). No subsequent saline flush of the IV line was allowed post-injection. To correct for attenuation in the PET data, a low dose computed tomography (CT) scan was performed. Brain images were acquired starting 90 minutes after the tracer injection, with six sequences of 5-minute frames over a duration of 30 minutes. For Aβ-PET imaging, the 18F-Florbetaben tracer was administered to all participants. The preparatory steps for these scans involved the insertion of an IV catheter, followed by a deliberate and gradual single IV bolus injection of 8.1 mCi ± 20% (300 MBq) of the tracer within 60 seconds or less (6 secs/mL max). Aβ-specific scans included two distinct post-injection imaging initiation times, with participants undergoing scanning 50 minutes after the tracer injection. To correct for attenuation, a low-dose CT scan was also performed. Brain images for each Aβ PET scan were captured in 4 sets of 5-minute frames over a span of 20 minutes.

### 2.3. Image preprocessing

We employed an in-house-developed fully automatic quantification method for PET scans, which was implemented and evaluated using histopathological data^28,30–33^. The process began by aligning dynamic PET frames to the first frame through rigid-body registration and then averaging them to generate a static PET image. This PET image was subsequently registered with the MRI and merged to create a composite image within the PET static space. Each individual’s MRI image in FreeSurfer^34^ (http://surfer.nmr.mgh.harvard.edu/) space was also registered to the same subject’s PET composite image using normalized mutual information and six degrees of freedom, resulting in a rigid-body transformation matrix. This matrix transferred all Schaefer atlas and FreeSurfer regional masks to the static PET image space. The standardized uptake value (SUV) was calculated for selected regions and then normalized to the cerebellum gray matter to derive the standardized uptake value ratio (SUVR). Building on our prior finding involving normative young subjects to identify Aβ abnormalities^28^, this study employed a global cut-off value (1.25) to identify Aβ-positive subjects. We specifically targeted regions of interest, encompassing the frontal, parietal, temporal, anterior, and posterior cingulate, as well as the precuneus regions.

Our rs-fMRI processing pipeline combines steps from the FSL toolbox^35^ and in-house-developed techniques^36^. To summarize, slice timing correction was applied to the raw rs-fMRI time series to account for the difference in the acquisition delay between slices. Simultaneously, motion parameters were estimated from the raw rs-fMRI scans using rigid-body registrations performed on all volumes in reference to the first volume. Subsequently, the estimated motion parameters and geometric distortion field were combined and applied to the slice-timing-corrected rs-fMRI time series to obtain undistorted and realigned rs-fMRI data. This matrix was constructed by calculating the correlation between the time series of the fMRI signals for all pairs of brain regions using Pearson’s correlation coefficient.

In this study, we employed the Schaefer atlas to parcellate the brain into 200 distinct regions and seven functional networks. Functional networks in the brain are cohesive groups of regions that collaborate during various cognitive processes (in the absence of any cognitive task). These seven functional networks are categorized as visual, somatomotor (referred to as motor in this study), dorsal attention, salience/ventral attention (referred to as salience in this study), control, limbic, and default mode networks. It is essential to note that the primary emphasis of our study’s results lies in the limbic network. This focus is driven by the observation that tau pathology tends to initiate within regions of the limbic system.

### 2.4. Subject categorization

In this study, we employed and expanded our previously published^28^ method to categorize older participants based on thresholds derived from the global Aβ and Braak stages I-II (entorhinal cortex and hippocampus) tau uptake levels observed in young participants. Measuring Braak stages I-II tau uptake was specifically employed to detect early abnormality stages. For Aβ, we computed the global SUVR by targeting specific regions of interest, including the frontal, parietal, temporal, anterior, and posterior cingulate, as well as the precuneus regions. We assessed the distribution of global Aβ and Braak stages I-II tau levels in the young participants. Subsequently, we computed the cut-point values for identifying abnormal global Aβ (cut-point=1.25) and Braak stages I-II tau (cut-point=1.08) by utilizing the 95th percentile of the fitted normal distribution. This categorization approach led to the classification of each participant into one of four groups: nAβ/nTau: Participants displaying neither abnormal global Aβ nor Braak stages I-II tau pathologies; aAβ/nTau: Participants characterized by normal Braak stages I-II tau levels but abnormal Aβ; nAβ/aTau: Participants exhibiting abnormal Braak stages I-II tau levels but normal Aβ pathologies; aAβ/aTau: Participants demonstrating abnormal Aβ and Braak stages I-II tau pathologies.

### 2.5. Graph theory

Graph theory serves as a foundational mathematical framework for examining graphs^37–39^, which are structures composed of nodes representing brain regions interconnected by edges signifying functional connectivity between these regions. We converted the undirected graphs into sparse ones by applying a threshold that retained the top 19% of the strongest weights^40,41^. We then computed the assortativity metric on the group average of the sparse functional connectivity map. The assortative measure assesses the tendency of nodes with similar degrees (i.e., the number of connections) to connect with others, and this can have a profound effect on the flow of information in the network ^42,43^. This measure varies from -1 to 1. The higher positive number illustrates the more assortative network which means the nodes with higher degrees are more likely to connect with other high-degree nodes.

### 2.6. Between-networks hyperconnectivity

In our longitudinal analyses, the between-networks hyperconnectivity metric quantifies the increased connectivity between functional networks, essentially measuring the level of integration between each network and others. To calculate the between-networks hyperconnectivity, we utilized the connectivity map at baseline and follow-up for each subject and network. For a more precise interpretation of hyperconnectivity, we excluded all anti-correlation coefficients (negative values) in both baseline and follow-up connectivity maps. Initially, we computed the difference between the follow-up and baseline connectivity maps for each subject, capturing subject-specific changes in connectivity patterns over time. To address yearly variations in each subject, we calculated changes in connectivity on an annual basis across subjects. Then, we measured the functional connectivity difference (correlation coefficient) between pairs of brain regions across different functional networks (between networks connectivity). In the final step, we determined the average of only the positive changes between functional network connectivity (increase in correlation coefficient in follow-up compared to baseline). This approach lets us to specifically focus on connections where connectivity has increased, providing a metric for hyperconnectivity. Consequently, we obtained seven between-networks hyperconnectivity metrics for each subject based on the seven functional networks of the Schaefer atlas.

### 2.7. Statistical analyses

We conducted a series of statistical tests to assess differences in demographic and cognitive variables among subjects. Continuous variables, such as age and cognitive test scores, were analyzed using t-tests and analyses of variance (ANOVAs)^44^. Categorical variables, such as gender were evaluated with chi-squared (χ2) tests.

In our preliminary analysis, we conducted a partial correlation analysis to explore the relationship between Aβ and tau levels across seven different functional networks. In another analysis, we used the regression analysis to test our hypothesis regarding the impact of Aβ on functional connectivity through compensatory mechanisms. We examined the effect of Aβ on limbic network functional connectivity. This analysis utilized a multiple regression model for each functional connection, assessing its association with Aβ deposition while controlling for age, gender, and regional tau levels. The model took the following form:

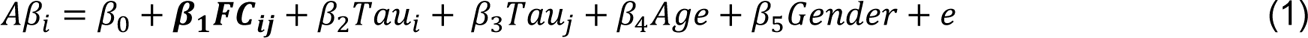

Where *i* varies between 1 to *N* (*i* ≠ *j*), and *N* is the maximum number of regions in the brain (across all seven networks); *j* represents the limbic network region and varies between 1 to 12 (*i* ≠ *j*) and 12 is a maximum number of regions in the limbic network. We implemented a multiple comparisons correction to account for family-wise error rates in regression analyses. This involved randomly shuffling the dependent variable 10,000 times to create a null distribution. Subsequently, we calculated the family-wise error rate-corrected t-value based on the 95th percentiles of the fitted normal distribution.

In longitudinal analyses, our primary goal was to evaluate the relationship between between-networks hyperconnectivity and the annual increases in Aβ and tau levels across all seven functional networks. Using regression analysis, we initially assessed the association between annual increases in Aβ and between-networks hyperconnectivity while controlling for baseline age, and gender. In separate regression analysis, we examined the association between annual increases in tau and between-networks hyperconnectivity.

Finally, we conducted mediation analyses to explore the role of between-networks hyperconnectivity within the limbic network in mediating the association between cortical Aβ in six spatially distinct networks (except the limbic network) and annual tau elevation within the limbic network. The total significance of the mediation effect was determined by the Sobel test^45^.

Our research used Python for all statistical analyses and data visualizations. Key numerical modules like NumPy and visualization tools like Matplotlib were pivotal in these analyses. For statistical tests, we relied on the SciPy statistical package (version 6.1.1)^46^. For graph theory analyses, we used the NetworkX package (version 3.2.1)^47^. In summary, our methodological approaches were comprehensive, encompassing a range of statistical analyses and controls to ensure the robustness and accuracy of our findings regarding the relationships between Aβ, tau, and functional connectivity in the brain.

## 3. RESULTS

### 3.1. Subject’s characteristics

In **Figures S1a-b**, we provide graphical representations of the regional distribution of tau using probabilistic atlases in both datasets, with regional cut-points determined based on normative young healthy subjects. **Table 1** offers comprehensive information about the number of subjects in each dataset, shedding light on their demographic profiles, pathological states, and assessment characteristics. There were significant differences observed in age (p-value < 0.0001), sex (p-value < 0.0001), race (p-value < 0.0001), and category fluency scores (p-value < 0.01) between the two datasets utilized in our study. As depicted in **Figure S1a**, the NOMEM dataset displayed distinctive tau abnormalities within the MTL since approximately 25% of subjects had tau that was observed outside of the temporal lobe. This dataset included 18% of subjects with a positive Aβ status. In contrast, within the BHII dataset (**Figure S1b**), tau pathology was predominantly limited to the MTL region. This observation suggests that subjects in this dataset are in the early stages of tau deposition, with tau pathology extending beyond the MTL in only a limited number of cases. Close to the NOMEM dataset, the BHII dataset included 23% of subjects with a positive Aβ status. However, the limited distribution of tau pathology and the percentage of subjects with tau abnormalities outside of the MTL are notably different in this dataset.

### 3.2. Association between Aβ and tau across different functional networks

In our initial exploration, we assessed the degree of correlation between Aβ and tau pathologies across various networks through partial correlation analyses. **Figure 1a** visually presents the partial correlation coefficients, highlighting these relationships across seven networks in the NOMEM dataset. To enhance our view of the correlation between Aβ and tau pathologies in each hemisphere of the brain, we separated the left and right hemispheres. Our findings revealed robust positive correlations, exceeding 0.5, between Aβ uptake in specific networks (including the bilateral salience, control, and default mode networks) and tau uptake in the left limbic network. This suggests a strong correlation between increased Aβ uptakes across these networks and increased tau uptake, particularly in the limbic network. The correlation between tau uptake in the left hemisphere of the limbic network and Aβ uptakes in other functional networks (both left and right hemispheres) is significantly higher than that of the right hemisphere (t-value = 4.81, p-value < 0.0001). Furthermore, we observed robust correlations, exceeding 0.48, between tau uptake in the left default mode network and Aβ uptake in various other networks (such as salience and control networks). This finding may be associated with the distribution of tau in this cohort, where several individuals exhibit tau outside the temporal lobe region.

**Figure 1.**
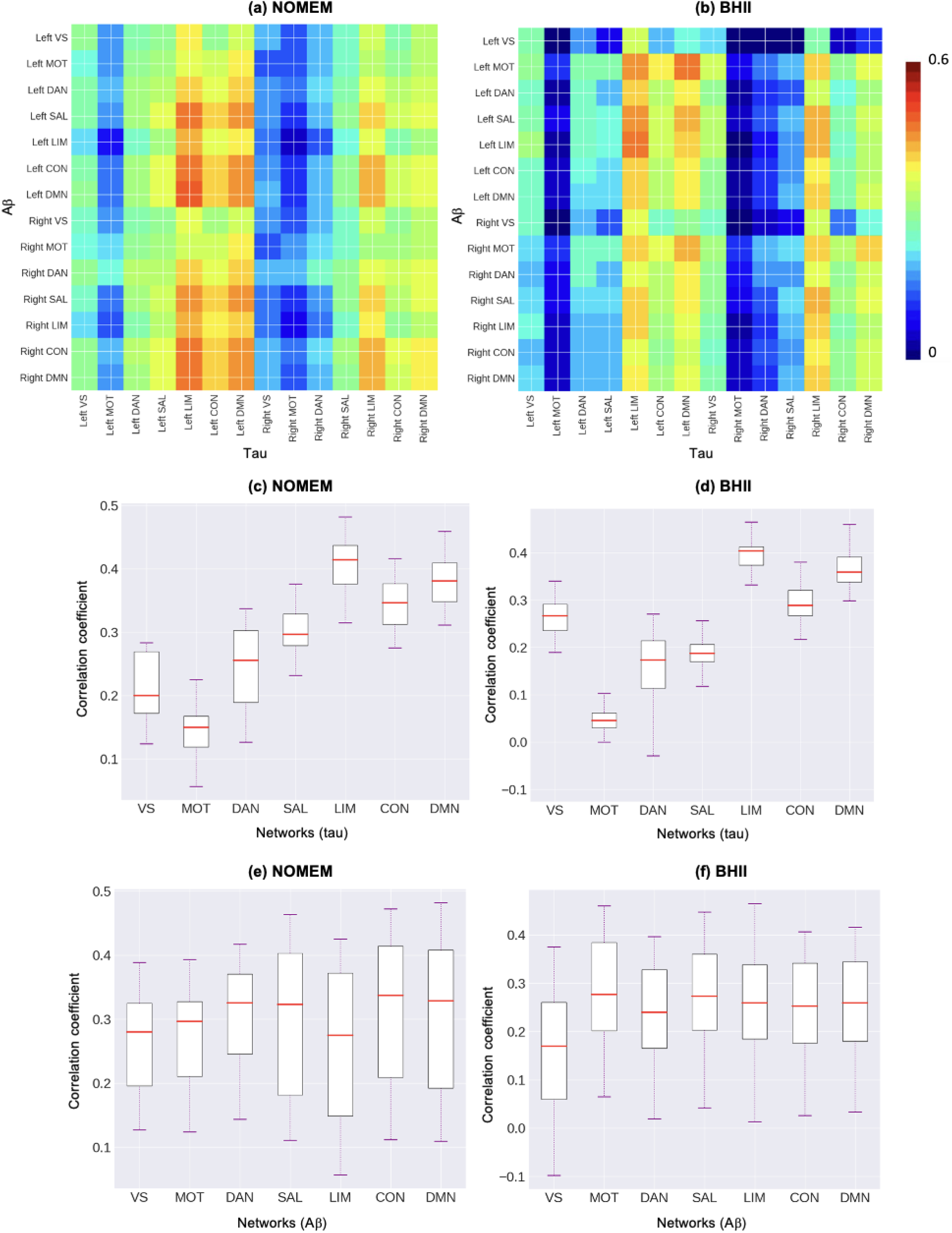
Internetwork partial correlation coefficients between Aβ and tau accumulations in seven functional networks in (a) NOMEM and (b) BHII datasets. This visualization depicts the correlation between Aβ and tau accumulations across different functional networks using a heatmap. The color scheme signifies the strength of the correlation, with red indicating a correlation value of 0.6 and higher and blue indicating a correlation value of 0. The boxplots compare the strength of the correlation coefficient of tau in seven different functional networks (x-axis) and Aβ in all the networks in (c) NOMEM and (d) BHII datasets. Additionally, boxplots compare the distribution of the correlation coefficient of Aβ in seven different functional networks (x-axis) and tau in all the networks in (e) NOMEM and (f) BHII datasets. VS: visual network, MOT: motor network, DAN: dorsal attention network, SAL: salience network, LIM: limbic network, CON: control network, and DMN: default mode network.

The findings from the BHII dataset in **Figure 1b** closely mirrored those obtained in the NOMEM dataset through the same analysis. Robust positive correlations (exceeding 0.44) consistently emerged between Aβ uptake in specific networks—such as salience, dorsal attention, and default mode networks—and tau uptakes in the left limbic network and left default mode network. Reiterating a consistent trend with the NOMEM dataset, the stronger correlation between tau uptake in the left hemisphere and Aβ uptakes across various networks was significantly higher than that of the right hemisphere (t-value = 2.11, p-value < 0.035). The results of the BHII dataset, underscore the robustness and reproducibility of our findings across different datasets.

To offer a comprehensive representation of the correlation matrix in **Figure 1a-b**, each column of the matrix was individually presented in **Figures 1c-d**. It is essential to note that the correlation coefficients from both the right and left hemispheres within each network were combined. This approach enabled us to depict the correlation of tau uptake in each network with Aβ uptakes across all networks. In **Figure 1c** (the NOMEM dataset), the limbic and default mode networks exhibited significantly higher correlation coefficients compared to other functional networks (t-value = 4.84, p-value < 0.0001). Also, in **Figure 1d** (BHII dataset), we observed that tau uptake in the limbic network displayed significantly higher correlation coefficients than the other six functional networks (t-value = 2.41, p-value < 0.019). This visual breakdown provides offers greater perspective of the distinctive patterns of association between tau and Aβ across all functional networks in both datasets.

We further separated each row of the correlation matrix (**Figure 1a-b**) into **Figures 1e-f**. It is essential to note that the correlation coefficients from both the right and left hemispheres within each network were combined. This approach lets us to visually represent the correlation of Aβ uptake in each network with tau uptakes across all networks. In both datasets (**Figures 1e-f**), all seven networks of Aβ exhibited competitive and relatively similar correlation coefficients with tau. These coefficients were not significantly different, underscoring the intricate nature of the relationship between cortical Aβ and tau across functional networks.

In summary, our results shed light on the complex relationships between Aβ and tau within different functional networks, emphasizing network-specific variations in the correlation of tau but not in Aβ. In other words, regardless of the spatial distribution of Aβ across functional networks, a noticeable correlation with tau is observed, particularly within the limbic and default networks.

### 3.3. Aβ increases the assortativity and connectivity between functional networks

We extended our analyses by categorizing subjects in both datasets into four groups: nAβ/nTau, aAβ/nTau, nAβ/aTau, and aAβ/aTau. This categorization enabled us to investigate how different combinations of Aβ and tau pathology influenced functional connectivity. Subsequently, we computed assortativity, a graph theory metric, on the average functional connectivity maps of each group (**Figure 2a-b**). Among the four subject groups in both datasets, aAβ/nTau subjects displayed the highest assortativity value (0.767 for NOMEM and 0.627 for BHII). Furthermore, within two subgroups of subjects with normal and abnormal levels of tau (nAβ/nTau versus aAβ/nTau and nAβ/aTau versus aAβ/aTau), the abnormality of Aβ levels consistently resulted in higher assortativity measures in both datasets. Higher assortativity values indicate a tendency for higher degree nodes to connect with other nodes possessing a higher degree of connection. This suggests that elevated Aβ levels may lead to increased connections with regions exhibiting a higher degree of connections (hub regions), even between functional networks.

**Figure 2.**
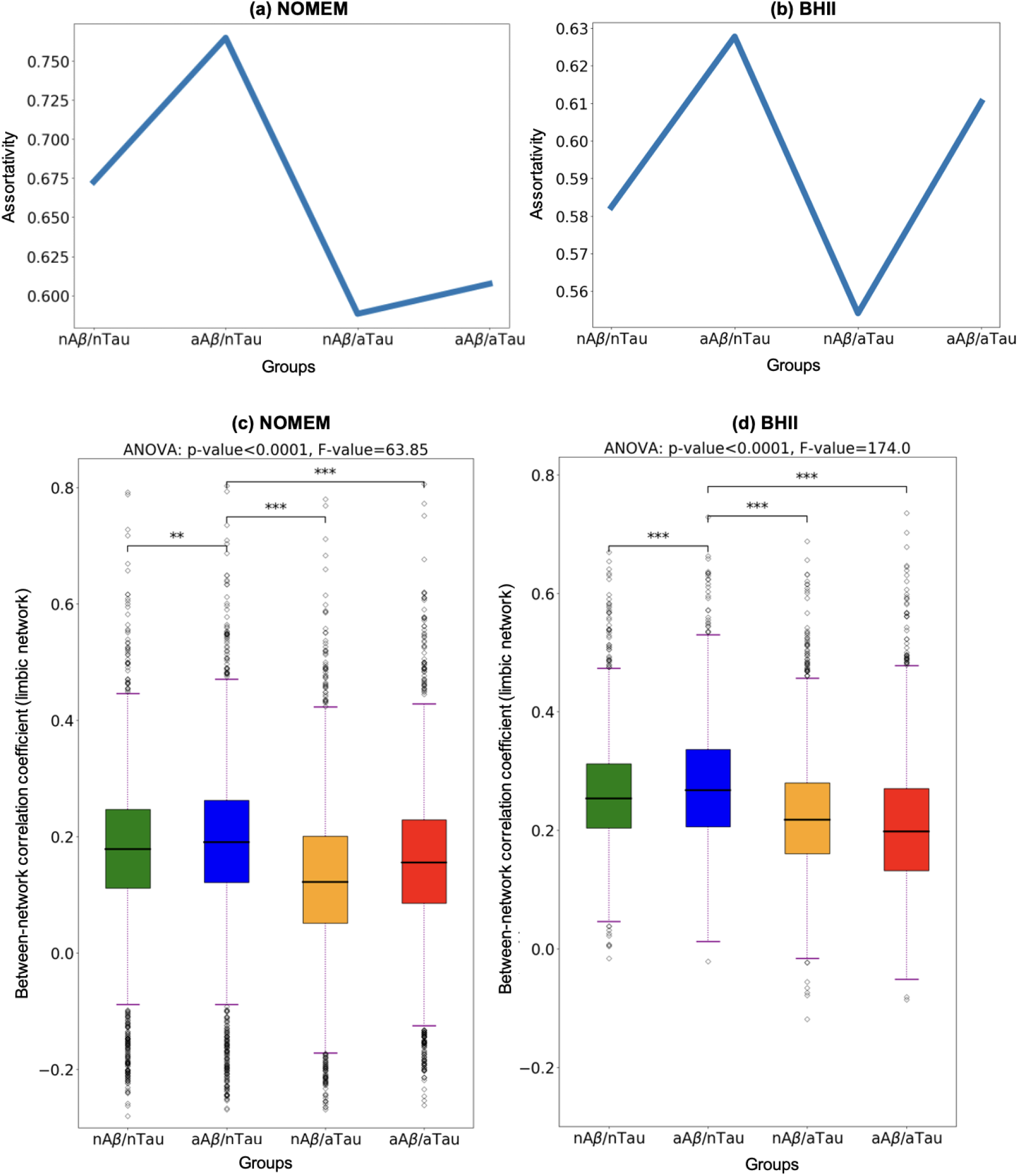
Functional network specification across four categories of subjects: nAβ/nTau, aAβ/nTau, nAβ/aTau, and aAβ/aTau. The assortativity measure was obtained from average functional connectivity in four groups in (a) NONMEM and (b) BHII datasets. Boxplots compare the distribution of between limbic network functional connectivity and other six networks correlation coefficients across four groups (nAβ/nTau, aAβ/nTau, nAβ/aTau, and aAβ/aTau) of subjects in (c) NONMEM (d) BHII datasets.

In **Figures 2c-d**, we conducted a comparison of between functional networks correlation coefficients involving the limbic network and the other six networks (i.e., visual, control, and default mode network). The objective was to distinguish variations in functional connectivity patterns across these four groups and explain the impact of cortical Aβ on limbic network connectivity with other networks. It is noteworthy that we did not exclude the anti-correlation coefficient (negative values) from connectivity map for these figures. **Figures 2c-d** show the results of this comparison in the NOMEM and BHII datasets, respectively. Our findings consistently indicate significant increases (t-value > 4.49, p-value < 0.0001) in between functional networks connectivity of the limbic network in the aAβ/nTau group compared with all other three groups of subjects (nAβ/nTau, nAβ/aTau, and aAβ/aTau) in both datasets. This increased between functional networks connectivity is particularly obvious in the presence of abnormal Aβ levels with normal levels of tau in the aAβ/nTau group. Thus, the presence of Aβ is associated with a potential compensatory response within the brain, manifested by an increase in connectivity between functional networks.

Moreover, **Figure S1a-b** visually presents the heightened connectivity within the limbic network (identified in yellow color) observed in the aAβ/nTau group compared to the nAβ/nTau group across both datasets. The edges in the figure specifically emphasize connections that exhibit a substantial increase, surpassing the threshold of 50%. This visual representation aims to elucidate the distinct patterns of increased limbic network connectivity, particularly with other functional networks. In **Figure S2a-b**, using all 481 subjects in both datasets and regression analyses (equation 1), we delved into the impact of Aβ on functional connectivity within the limbic network regions (identified in yellow color) across both datasets. **Figure S2a-b** visually show connections that displayed a significant positive association between Aβ uptakes in various functional networks and functional connectivity within the limbic network regions. The thickness of these depicted connections signifies their statistical significance (t-value > 2.10, p-value < 0.018 for NOMEM and t-value > 2.22, p-value < 0.014 for BHII) after corrections for multiple comparisons. Importantly, considering both datasets, the results illustrate the existence of at least one functional connectivity between the limbic network and all other functional networks, each demonstrating a significant association.

### 3.4. Associations between elevations of Aβ and tau with between-networks hyperconnectivity

We conducted longitudinal analyses in a cohort of 46 HC subjects. In **Figures 3a-b**, we investigated the association between change in between-networks hyperconnectivity and the annual elevations of Aβ and tau pathologies. When considering all seven networks, our analyses revealed a significant and noteworthy relationship. This relationship indicated a strong, significant association (t-value = 8.39, p-value < 0.0001) between an increase in elevation of Aβ and an increase in between-networks hyperconnectivity (**Figure 3a**). Subsequently as illustrated in **Figure 3b**, there is a significant association (t-value = 6.37, p-value < 0.0001) between an increase in between-networks hyperconnectivity and an increase in elevation of tau. In other words, higher levels of between-networks hyperconnectivity (attributed to an increase in annual Aβ based on this study’s hypothesis) were linked to an increase in the future levels of tau pathology.

**Figure 3.**
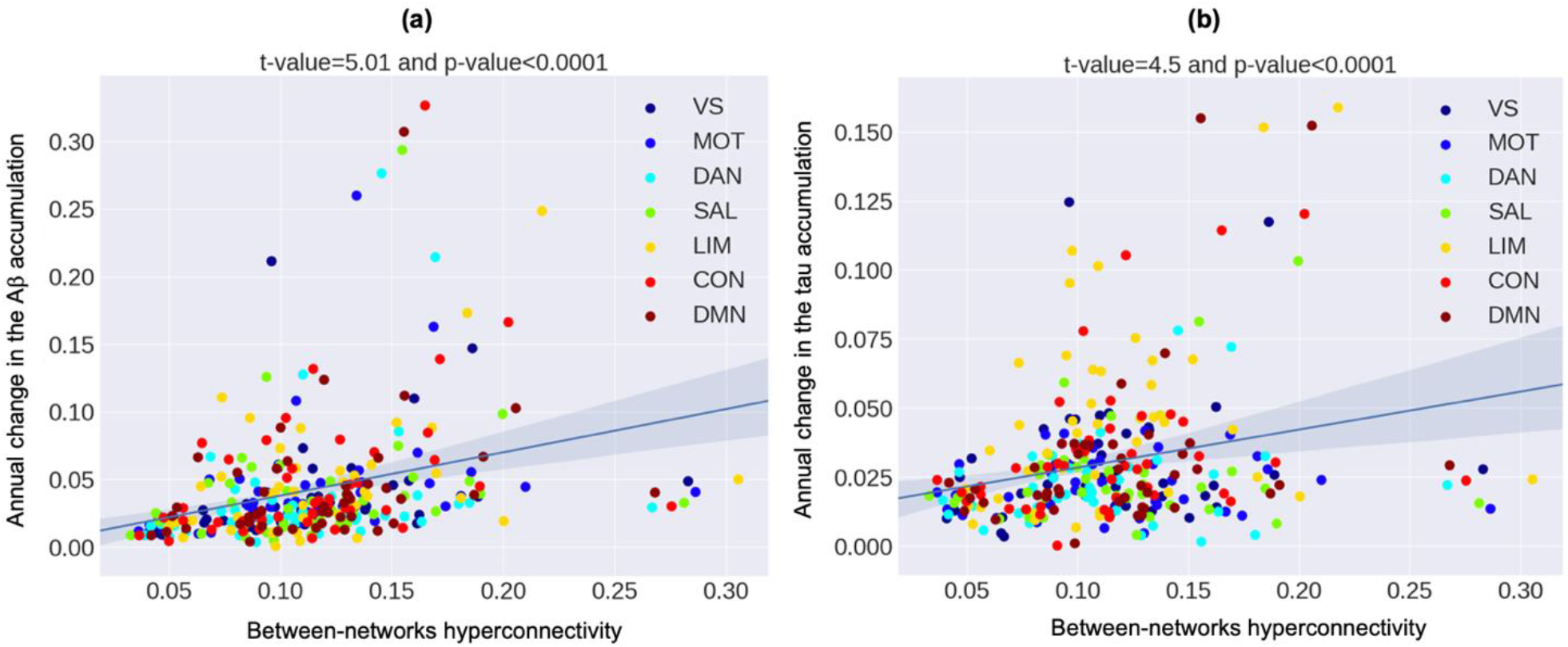
Associations between the annual change in (a) Aβ (b) tau levels with between-networks hyperconnectivity across all seven networks. VS: visual network, MOT: motor network, DAN: dorsal attention network, SAL: salience network, LIM: limbic network, CON: control network, and DMN: default mode network.

### 3.5. Between-networks hyperconnectivity of the limbic network mediates the associations between cortical Aβ and tau within the limbic network

Finally, we conducted mediation analyses to strengthen the evidence for our hypothesis, as depicted in **Figure 4a-f**. These mediation analyses aimed to explore the effects of the annual elevation of Aβ within six brain networks (default mode, control, dorsal attention, salience, visual, and motor networks) on annual tau changes within the limbic network, mediated by changes in between-networks hyperconnectivity of the limbic network. While the mediation analyses did not yield significant results in the entire longitudinal population of 46 subjects and the 32 Aβ-negative subjects (baseline global Aβ<1.25), they achieved highly significant results (Sobel test p-value < 0.013) for indirect relationships in all six networks for the 14 Aβ-positive subjects. The direct relationship between Aβ and tau was nonsignificant across all six networks. These results provide robust evidence supporting our hypothesis, indicating that the interplay between higher Aβ levels and increased between-networks hyperconnectivity serves as a strong mediator of the likelihood of tau elevation in the limbic network over time. This valuable insight sheds light on the key mechanisms underpinning AD progression, paving the way for a clearer view of the disease’s trajectory.

**Figure 4.**
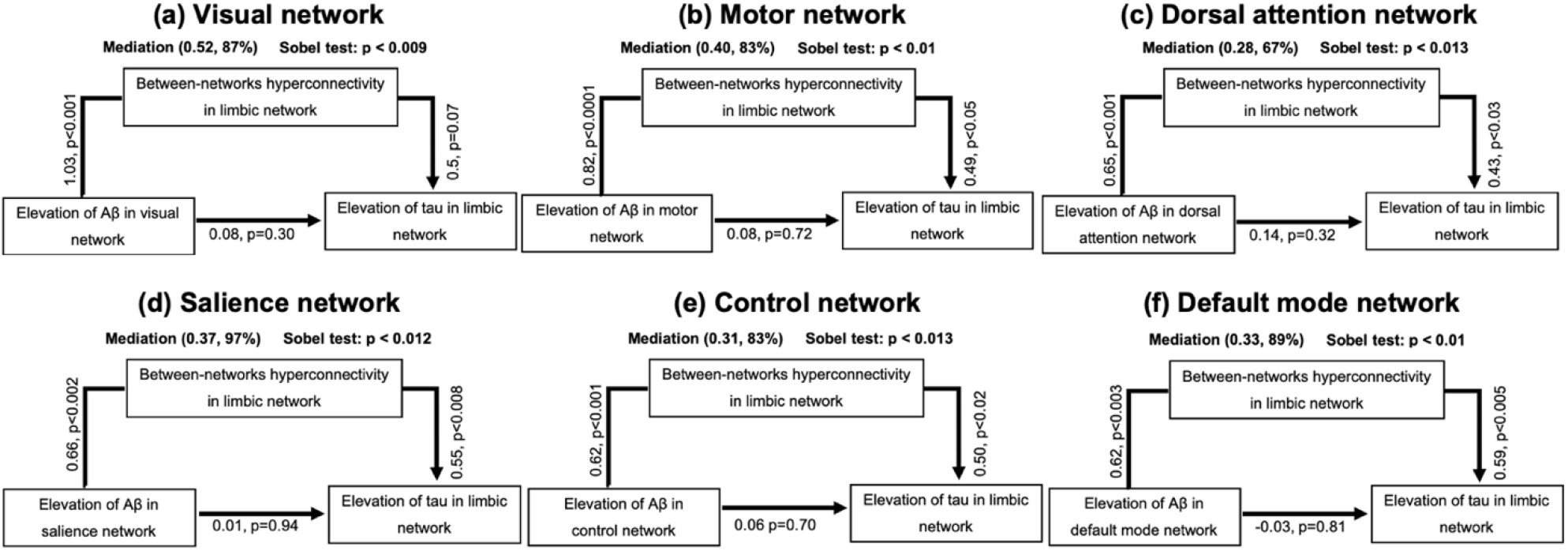
Role of between-networks hyperconnectivity of limbic network in mediating the association between elevations of cortical Aβ across six networks and limbic network tau elevation in (a) control network (b) dorsal attention network (c) default mode network (d) limbic network (e) motor network (f) visual network (g) salience network, and tau elevation within the limbic network.

The findings of this study provide valuable insights into several pivotal stages that illuminate the intricate relationship between AD pathologies and functional connectivity based on our hypotheses (as shown in **Figure 5**). These critical stages offer a window into the way these pathologies influence the brain’s connectivity patterns and ultimately contribute to the disease’s progression. The first crucial turning point, which marks the transition from normal aging to the early stages of AD, occurs when Aβ accumulates in neocortical regions. The accumulation results in hyperconnectivity between specific functional connections in the brain and may trigger compensatory mechanisms. These mechanisms represent the brain’s adaptive response to the increasing presence of Aβ, resulting in between-networks hyperconnectivity and functional network integration. Therefore, Aβ induces between-networks hyperconnectivity between distinct functional networks and limbic regions. The second critical stage, as depicted in **Figure 5**, represents a key phase where this between-networks hyperconnectivity mediates an early interplay between Aβ and tau pathologies within the MTL region. This mediation is pivotal in the disease process and underscores the complex dynamics between these two pathologies in the early stage of tau accumulation. The third critical stage arises when tau pathology initiates hypoconnectivity in response to the disease process. This phase marks a period where the relationship between Aβ and tau becomes intricate. Aβ promotes hyperconnectivity as a compensatory response to pathological changes, while tau’s presence contributes to hypoconnectivity. This balance between hyperconnectivity and hypoconnectivity eventually tips the scales with the spreading of tau, leading to more pronounced hypoconnectivity and cognitive deficits in the fourth critical stage.

**Figure 5.**
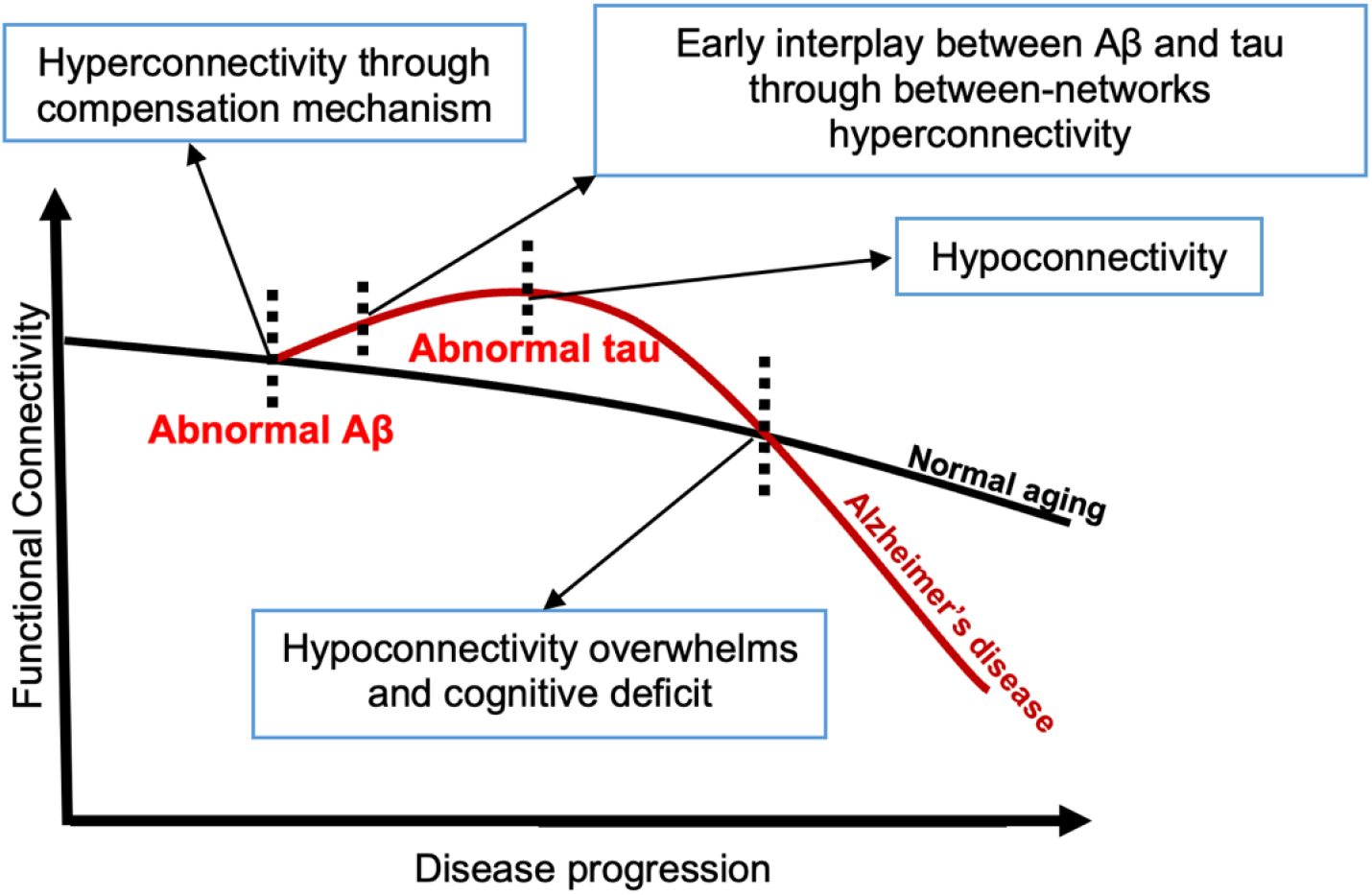
Proposed mechanism of functional connectivity role in the remote association between early-stage tau and cortical Aβ.

## 4. DISCUSSION

This research aimed to address two pivotal questions with far-reaching implications in AD pathogenesis: 1) whether Aβ accumulation triggers a compensatory response, leading to hyperconnectivity between networks and increased functional networks integration, and 2) whether this compensatory mechanism promotes further accumulation of tau. First, our observations indicated that during the early stages of accumulation, the limbic network exhibits the highest correlation to cortical Aβ in both datasets. This observation sheds light on the brain regions most affected by Aβ pathology during the initial phases of pathological progression. Second, our research revealed a compelling mechanism in which early Aβ accumulation triggers compensatory responses within the brain. This leads to hyperconnectivity and increased network integration, presumably as an adaptive strategy to counteract the disruption caused by Aβ. Finally, we demonstrated the mediation effect of between-networks hyperconnectivity on remote association between elevations of Aβ and tau in distinct functional networks. This mediation effect of neural functioning adds a valuable dimension to our understanding of disease progression, potentially informing early intervention strategies in AD.

It has been observed that the association between Aβ and tau pathologies often initiates when there is cortical Aβ deposition and tau starts to deposit in the limbic network, especially in the entorhinal cortex, which is one of the earliest regions of tau accumulation^29,48^. This interaction between Aβ and tau in the limbic network is believed to initiate the spread of tau pathology to other brain regions through network connections. This spreading of tau is thought to underlie the progression of cognitive deficits in AD. Previous research has shown that the superficial layers like layer II/III, are more vulnerable to AD pathology, including Aβ and tau accumulation^49^. Since the superficial layers are more involved in corticocortical connections ^50^, this vulnerability is related to the neural connections and circuits within the brain ^49^. In other words, AD pathologies are more likely to affect regions with extensive connections to other brain areas. Therefore, it’s reasonable to suggest that in the early stages of pathologies, the differences in vulnerability to pathological elevations arise from the neural pathways, particularly functional connections, rather than the colocalization of Aβ plaques with tau tangles occurring in later stages.

In our previous study^33^, we investigated the unique impact of Aβ and tau proteins by categorizing them into normal and abnormal groups. Our findings unveiled compelling insights into the early stages of Aβ deposition (with normal levels of Aβ), showing a notable association with increased cortical thickness, particularly in the MTL region. These observations provide evidence for a compensatory mechanism within the brain when faced with Aβ plaque accumulation. In other words, altering the balance between synaptic excitation and inhibition^1^ can lead to increased neural activity, potentially inducing cortical thickening and contributing to the onset of neuroinflammation. Building upon this research, our investigations were extended to include the association between these two pathologies across spatially remote (not spatially overlapped) regions^29^. This study uncovered a strong association between tau in the MTL and Aβ in different cortical brain regions in the early stages of tau progression. This evidence could be attributed to the enhanced possibility of neural connections between Aβ and tau from distant brain regions. Previous research also has reported increased pathology-associated posterior default mode network connectivity in individuals with low Aβ levels ^51,52^. These observations collectively underscore our hypothesis that the accumulation and spread of pathological tau are intricately intertwined with cortical Aβ pathology through neural function and the interconnectedness of brain regions.

Despite the complex relationship between AD pathologies and functional connectivity, we took a conservative approach to control each pathology to examine its association with the other, along with functional connectivity. While much research has focused on the impact of Aβ on within-network connectivity^17,53^, in this study, we found that Aβ levels are highly associated with between-networks hyperconnectivity. This broader perspective is crucial because cognitive functions often rely on the collaboration of multiple brain networks rather than the isolated functioning of individual networks^54^. Specifically, a noticeable shift in the balance between functional networks has been observed as individuals age^20,21^. This shift is reflected in a decline in modularity, a measure of how distinct and specialized functional brain networks are. Moreover, it has been shown that higher-level functions, such as executive and perceptual processing, may decline with age^55,56^, while basic information processing, like primary sensory and motor functions, remains relatively intact^22^. This suggests that higher-level functions may also need to recruit more neural resources further down the processing stream. Collectively, these findings illustrate the nature of brain connectivity as individuals age, where the allocation of neural resources and an increase in connectivity with specific networks (compensation mechanism of the brain) can play significant roles in cognitive processes and response to pathological changes during the progression of the disease.

The selective optimization with compensation theory has supported compensatory mechanisms in the brain, which suggests that as individuals age, there is a decline in the separation or segregation of brain regions ^57^. However, in this study, we provide robust evidence that the compensatory response is triggered by Aβ, which increases functional connectivity and establishes pathways connecting the limbic network to other functional networks. This compensatory response to abnormal Aβ can be explained by several biological hypotheses. Abnormal Aβ may involve reinforcing the structure of synaptic connections in the brain, which is associated with an increase in synaptic activity^58^. This synaptic enhancement may be facilitated by the generation of new neurons through neurogenesis^59,60^, helping to reintroduce fresh neurons into the disrupted neural network. This compensatory mechanism may also be triggered by the rising of brain metabolism^61,62^ followed by protective antioxidant activity^63,64^ reported by several previous studies to counteract the damage in neural functioning. Finally, current reports consistently observe activated microglia at sites where aggregated Aβ has accumulated, indicating a potential role for microglia in clearing brain Aβ ^65^ and early improvements in synaptic plasticity through increased neurotransmitter release^66,67^. This compensatory response by microglial activation could potentially lead to hypoconnectivity and hyperactivation, which enhance the connections between different functional networks.

According to the *amyloid cascade hypothesis*^68^, cortical Aβ facilitates the initiation or exacerbation tau-related abnormalities further down the line. Recent studies have suggested a pivotal theory that Aβ plays a facilitating role in the spread of tau beyond the MTL. The lack of a mechanism to explain this remote association of Aβ on early-stage tau has led to various studies opposing the *amyloid cascade hypothesis* and reporting the independent progression of early-stage tau and Aβ in several studies^69–71^. In this study, we have provided evidence supporting our main hypothesis that the interaction of Aβ and tau mediates by between-networks connectivity. To our knowledge, no other study has demonstrated this mediation effect thus far. However, previous studies illustrated the significant link between neural activity and Aβ deposition^72,73^, leading to changes in activation patterns in the MTL region^19,74,75^. On the other hand, in vivo, research has reported that tau pathology spreads through trans-synaptic networks, potentially related to broader network disruptions in long-distance brain regions through neuronal projections^26,76,77^. So, the trans-synaptic spread of tau may act as a bridge explaining the broader associations of Aβ in remote regions through neuronal projections in functionally connected regions. These findings highlight the dynamic nature of brain network organization and its ability to adapt in response to cognitive challenges, potentially serving as a compensatory response to Aβ pathology and ultimately leading to elevation of tau accumulation and spreading. The findings generally enhance our knowledge of AD pathogenesis and offer new paths for research and potential therapeutic interventions targeting this devastating disease.

It’s important to acknowledge and address certain potential limitations in this study, which should guide future research endeavors. Firstly, this study employed HC elderly subjects using the second generation of the tau tracer (F18-MK6240), resulting in a relatively limited sample size with abnormal Aβ and tau. This limitation may have impacted the statistical power of certain analyses, such as those involving four group categorization and longitudinal cohorts. However, it’s noteworthy that previous studies have also worked with cohorts of similar or even smaller sizes. To ensure the robustness of our findings, the results of this study must be verified in independent datasets with a larger longitudinal sample size. Another limitation pertains to the BHII dataset, which primarily consists of individuals in earlier stages of AD, potentially representing a different pathological stage compared to that of the NONMEM study. This limitation prevented us from fully replicating the findings related to the relationship between Aβ and functional connectivity. Nevertheless, we have conducted a variety of analyses to provide support for our hypotheses. Furthermore, it’s important to highlight the absence of genotype information in this study population, particularly information regarding APOE status, which could serve as a valuable covariate in our regression analysis. While these limitations exist, they should be viewed as opportunities for further research to build upon and enhance our understanding of the complex relationships between tau, Aβ, and functional connectivity in AD.

## 5. CONCLUSION

In this study, we have highlighted the early accumulation of Aβ and its role in triggering compensatory mechanisms. This compensatory response enhances integration between functional networks, adopting between-networks hyperconnectivity. Subsequently, the between-networks hyperconnectivity mediates the remote relationships between Aβ and tau pathologies in the early stages of the disease. Notably, the between-networks hyperconnectivity has emerged as a pivotal mediator in the relationship between early-stage tau in limbic network and Aβ in distinct functional networks. The study reveals crucial aspects of AD pathogenesis, paving the way for a more profound understanding of AD progression and potential interventions in the early stages of the disease.

## Data Availability

All data produced in the present study are available upon reasonable request to the authors

## Ethical consideration

Our data and any subsequent studies derived from it have undergone thorough ethical approval through the processes of two Institutional Review Boards (IRBs), located at Columbia University and Weill Cornell Medicine. These regulatory bodies play a crucial role in ensuring the integrity of our research by safeguarding the rights, well-being, and confidentiality of research participants.

The approval from IRBs signifies that our research design, methodology, and associated procedures adhere to stringent ethical standards and regulatory guidelines. This comprehensive review process assures that our research is conducted with the utmost consideration for ethical principles, addressing critical factors such as participant consent, data confidentiality, and the overall welfare of individuals involved in the study. This approval underscores our commitment to ethical research practices and reinforces the credibility and reliability of our data and subsequent studies.

## Supplementary materials

**Table S1.**
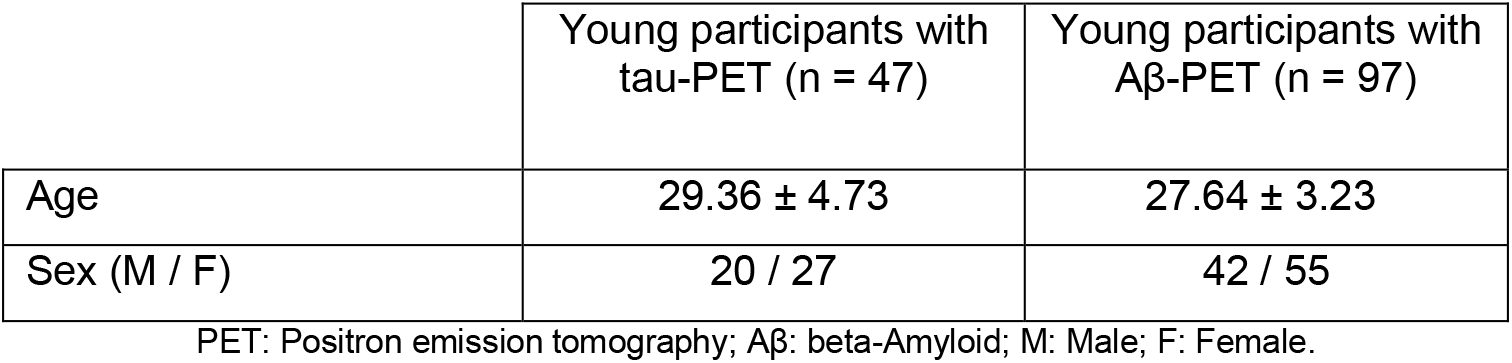
Demographics of the young subjects.

**Figure S1.**
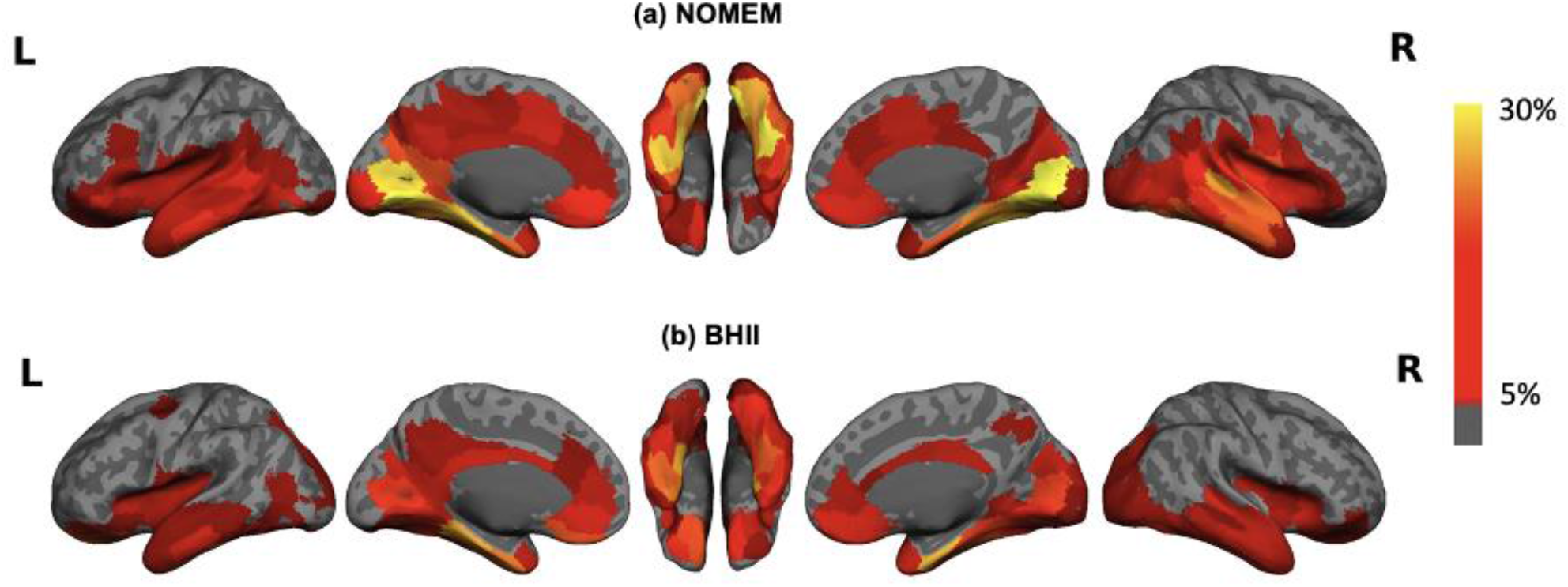
A visual representation of region-wise probabilistic atlas depicting the spatial distribution of tau pathology across the entire cerebral cortex in (a) NOMEM and (b) BHII datasets. We used a color-coded heat map to represent the probability of observing tau at each region, compared to a young control cohort, which is then overlaid onto a semi-inflated cortical surface of the MNI152 template.

**Figure S2.**
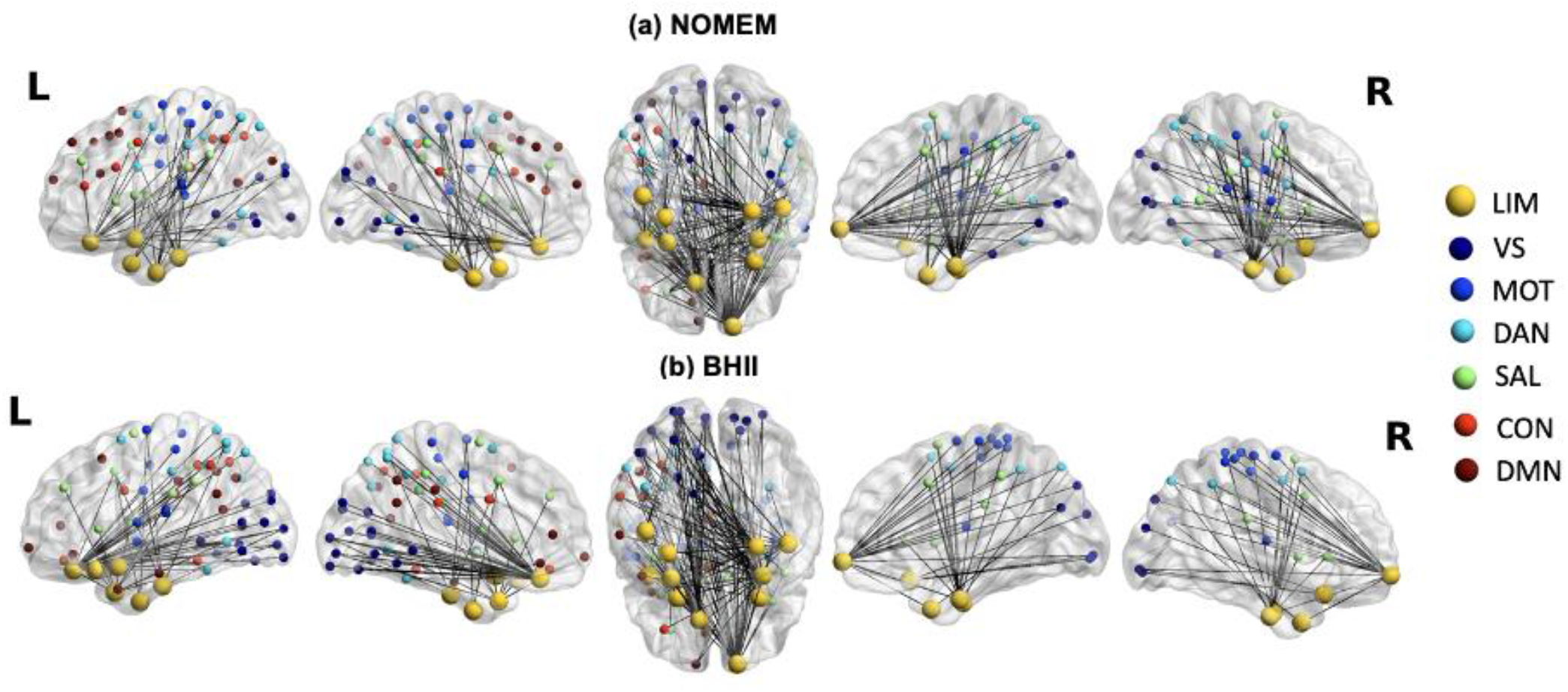
Illustrating the increase in limbic network functional connectivity of aAβ/nTau compared with nAβ/nTau in (a) NONMEM and (b) BHII datasets. The edges correspond to the connections that displayed differences of 50% or higher. Functional networks are color-coded and overlaid onto the semi-inflated cortical surface of the MNI152 template particularly the limbic network brain regions represented with yellow color with bigger size of nodes than other six different networks. VS: visual network, MOT: motor network, DAN: dorsal attention network, SAL: salience network, LIM: limbic network, CON: control network, and DMN: default mode network.

**Figure S3.**
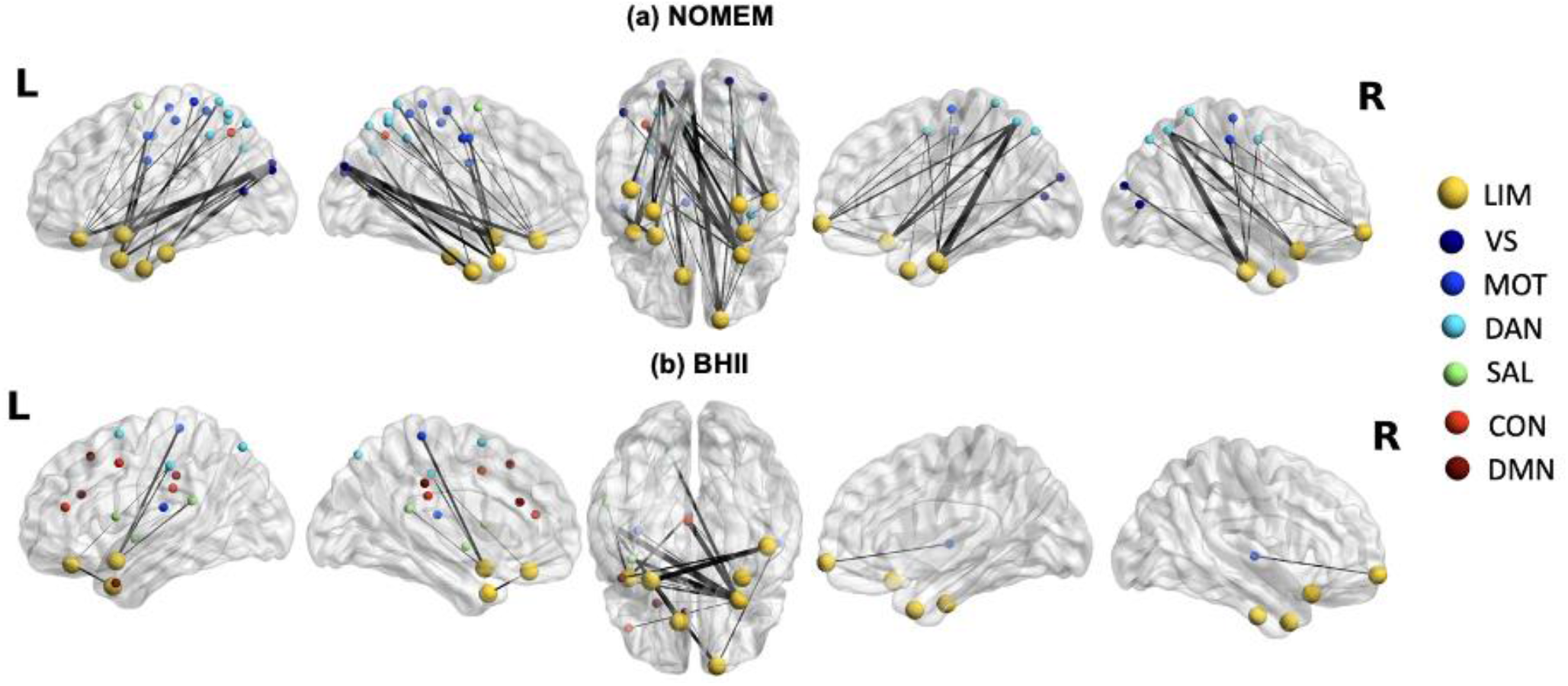
Statistical connectivity maps (t-values) illustrating the associations between regional Aβ and functional connectivity within the limbic network in (a) the NOMEM and (b) the BHII datasets. The edges between nodes (brain regions across seven networks) signify the connections from limbic network regions to cortical Aβ accumulation, and the thickness of these edges corresponds to the level of statistical significance (t-value). Functional networks are color-coded and overlaid onto the semi-inflated cortical surface of the MNI152 template particularly the limbic network brain regions represented with yellow color with bigger size of nodes than other six different networks. Only associations that survived multiple comparison corrections are displayed. VS: visual network, MOT: motor network, DAN: dorsal attention network, SAL: salience network, LIM: limbic network, CON: control network, and DMN: default mode network.

